# REINA: A Recognize-Then-Infer Wearable-to-App AI Framework for Breast Cancer Rehabilitation

**DOI:** 10.64898/2026.08.29.26361725

**Authors:** Qiao Zhuang, Changhong Mou, Bowen Liu, Gregory King, Mei Rosemary Fu

## Abstract

Breast cancer survivors frequently experience upper-limb impairments, making continuous monitoring essential for effective rehabilitation. We propose REINA (Recognize-Then-Infer Wearable-to-App <u>A</u>I Framework), a two-stage deep-learning approach for remote monitoring of motor function during breast cancer rehabilitation using wearable-device data. Inertial measurement unit (IMU) signals from wearable devices are first used to recognize physical activities via supervised learning, followed by an activity-specific recurrent neural network (RNN) to infer corresponding electromyography (EMG) signals. REINA establishes reliable inference of neuromuscular activity from wearable IMU data, enabling real-time, cost-effective assessment of motor function recovery in real-world settings.

## 1 Introduction

Breast cancer survivors frequently face upper-limb impairments, lymphatic pain, and lymphedema, making continuous rehabilitation critical, as recovery heavily relies on regaining balanced and functional use of the arms in daily life [1, 2, 3]. Tracking muscle recovery and functional movement requires electromyography (EMG) [4]. However, EMG assessments can be costly [5, 6], demand specialized equipment alongside trained personnel, and are ultimately not feasible for routine or long-term monitoring outside of clinical settings. Consequently, clinic-based assessments provide only a limited snapshot of a patient’s true physical capability and rehabilitation progress [7].

To overcome these limitations, continuous observation in real-world settings is necessary [8]. Wearable technology, such as unobtrusive inertial measurement units (IMUs) and smartwatches, presents a practical solution for this, enabling the continuous capture of upper-limb kinematics during daily activities. However, relying on these devices introduces a new scientific challenge: a significant gap exists between the broad movement patterns recorded by wearable devices and the precise, localized muscle activation traditionally measured by EMG. *Establishing a robust, quantifiable connection to determine how wearable signals can accurately reflect hidden neuromuscular function is a critical scientific question*. Bridging this gap is essential for turning everyday sensor data into the rigorous, clinically meaningful metrics of muscle recovery necessary for effective rehabilitation. Mathematically, this requires discovering a *mapping* between two distinct spaces: the lower-dimensional kinematic manifold captured by wearables and the high-dimensional, highly non-linear manifold of neuromuscular activation. Because the overactuated musculoskeletal system allows multiple distinct neuromuscular activation strategies to generate the exact same time-parameterized kinematic trajectory [9, 10], establishing a direct, one-to-one regression mapping between these spaces is mathematically unavailable. Consequently, recovering the hidden physiological states from observable kinematic data constitutes a highly complex, ill-posed inverse problem governed by underlying biomechanical dynamics.

Developing a deep-learning-driven musculoskeletal digital twin (DT) offers a transformative approach to solving this challenge [11, 12]. To overcome the aforementioned mathematical challenge of a non-injective mapping, such DT could potentially employ Recurrent Neural Networks (RNNs). Because human motor control exhibits strong non-Markovian properties, where current neuromuscular activation relies heavily on the continuous historical trajectory of the movement rather than just the immediate kinematic state, the RNN effectively resolves this ill-posed inverse problem by capturing these essential temporal dependencies. By successfully translating everyday wearable data into actionable physiological insights, this robust capability allows the DT to serve as a highly applicable medical tool. It empowers clinicians to continuously monitor remote rehabilitation progress, precisely identify functional asymmetries during a patient’s daily life, and ultimately drive highly personalized, data-driven therapeutic strategies outside the traditional clinical setting.

In this work, we propose a Recognize-Then-Infer Wearable-to-App <u>A</u>I Framework for Breast Cancer Rehabilitation (REINA). REINA is a two-stage, deep-learning-driven framework designed to provide a practical and low-cost method for assessing breast cancer rehabilitation outside the clinic. To successfully map wearable-derived kinematics to hidden physiological states, the proposed architecture employs a “Recognize-then-Infer” methodology consisting of two primary steps. First, wearable motion signals, specifically continuous accelerometer and gyroscope time series from Inertial Measurement Units (IMUs), are utilized to recognize the physical activity being performed. This step operates on the premise that, while the recorded kinematic data are inherently complex, signals generated by different physical activities reside on distinct, activity-specific kinematic manifolds. Given the highly nonlinear and dynamic topology of these spaces, standard classification methods often fall short [13, 14]. Therefore, we leverage supervised deep learning techniques, which possess the representational capacity needed to disentangle these complex manifold structures and separate the various movement patterns. Second, once the specific activity is identified, the same wearable time-series signals are fed into an activity-specific inference model to estimate the corresponding EMG signals. In this stage, we employ a recurrent neural network (RNN) to capture the critical temporal dependencies between the smartwatch measurements and neuromuscular activity. Because muscle activation at any given moment is governed not only by the current motion but also by the recent movement history, the RNN effectively resolves this sequence-to-sequence mapping. By accurately inferring muscle activity directly from accessible wearable motion data, this framework provides a mechanism to assess whether patients are executing rehabilitation movements safely and appropriately. Finally, our two-stage REINA framework provides a novel and applicable tool for breast cancer rehabilitation. By first identifying the patient’s movement and then estimating the underlying muscle activity, this framework seamlessly bridges the gap between everyday wearable and clinical biomechanics. It allows for continuous monitoring of recovery in real-world settings, helping clinicians identify functional differences between limbs and customize rehabilitation strategies to each patient.

The remainder of this paper is organized as follows. Section 2 details the methodology of the proposed REINA framework, including multi-window aggregation in a supervised learning framework for wearable motion classification and the subsequent RNN inference for breast cancer rehabilitation. Section 3 presents the data and results, covering the data sources, motion trajectory point clouds of the participants, and the predictive performance of both the supervised-learning classification and RNN inference model. Finally, Section 4 summarizes the conclusion and outlines directions for future work.

## 2 Methodology: REINA Framework

The goal is to provide a practical and low-burden way to assess functional recovery outside the clinic, where traditional monitoring tools such as electromyography (EMG) are costly and seldom available for clinical use. Additional EMG monitoring requires specialized equipment and trained personnel, and are often not feasible for routine and ongoing or long-term use [15, 16]. The proposed framework consists of two main steps that are also illustrated in Figure 1.

**Figure 1:**
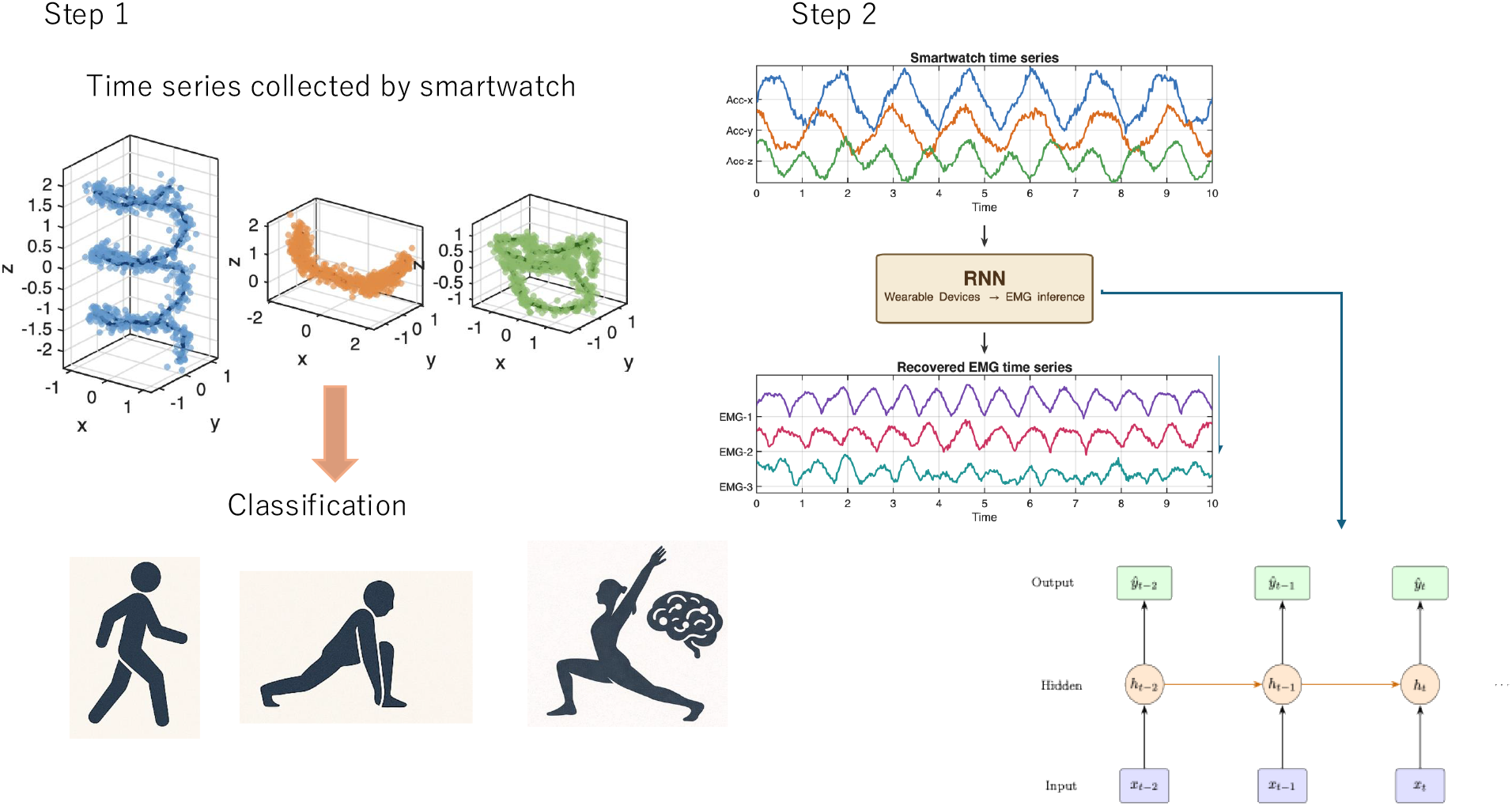
Illustration of the proposed REINA framework

**Figure 2:**
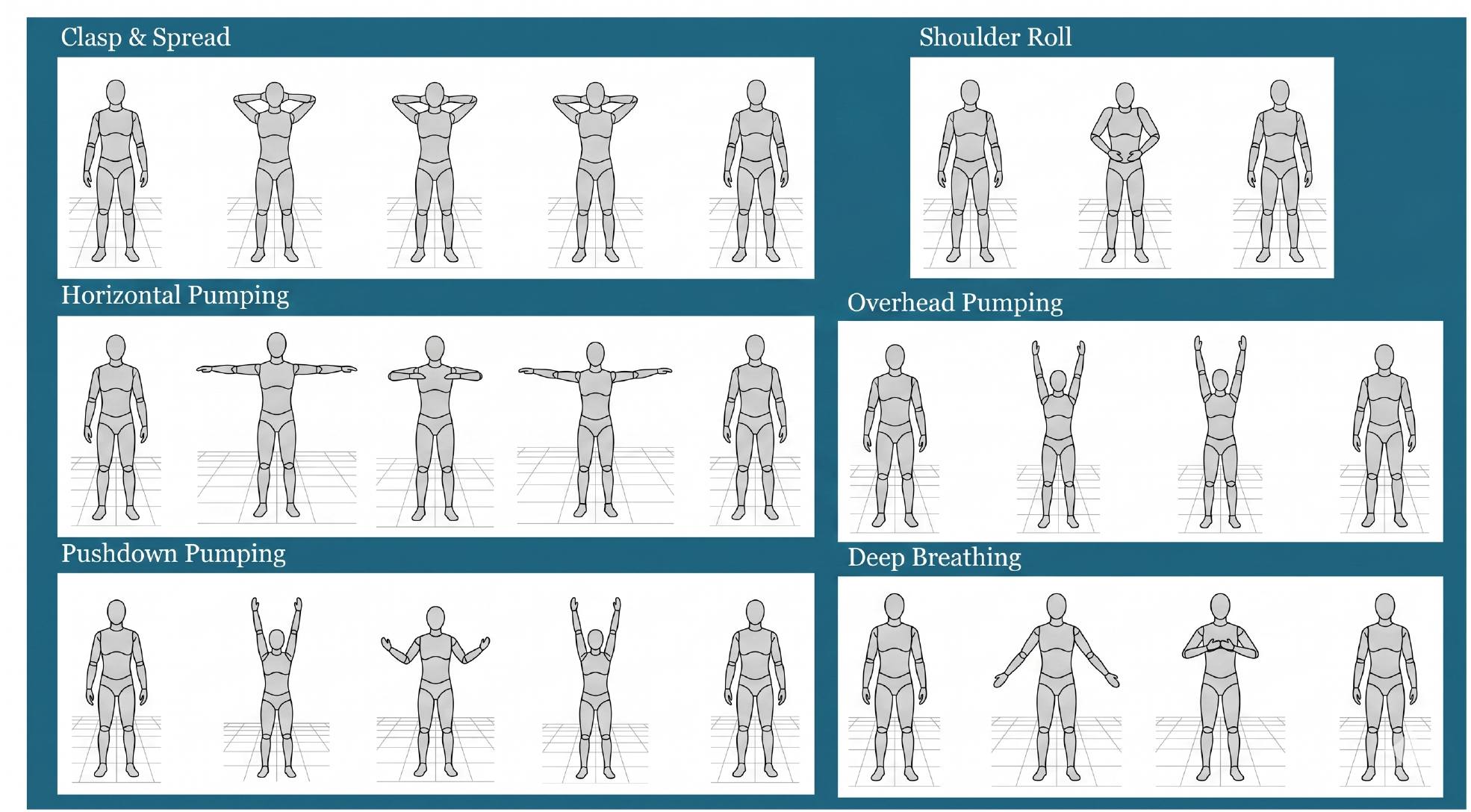
Standardized The-Optimal-Lymph-Flow (TOLF) upper-limb exercises performed by all participants.

First, smartwatch-based motion signals, including gyroscope and accelerometer time series, are used to recognize the physical activity being performed. In this step, we assume that motion signals generated by similar activities share an underlying low-dimensional structure, even though the recorded data are complex and time-dependent. Based on this idea, data-driven methods are used to identify shared movement patterns and group similar motions together [17, 18]. This provides the activity context for the next stage and helps distinguish among different rehabilitation-related movements. Second, once the activity is identified, the same wearable time-series signals are used in an activity-specific inference model to estimate the corresponding EMG signals. Here, we use a recurrent neural network (RNN) to capture the temporal dependence between recent smartwatch measurements and muscle activity, since muscle activation at a given moment is influenced not only by the current motion but also by the recent movement history. By inferring muscle activity from wearable motion data, the framework can help assess whether breast cancer survivors are performing rehabilitation movements in a safe and appropriate way.

This approach is particularly relevant for breast cancer rehabilitation, where recovery often depends on regaining balanced and functional use of the upper extremities in daily life. Clinic-based assessments provide only a limited snapshot, whereas wearable devices make it possible to observe movement continuously in real-world settings [19]. By combining activity recognition with muscle activity estimation, the proposed method can help track rehabilitation progress, identify differences between the affected and non-affected sides, and support more personalized rehabilitation strategies.

### Summary of the notations

For ease of reference, the main symbols and notations used throughout the paper are summarized in Table 1.

**Table 1:** Table of Notations.

| Notation | Description |
| --- | --- |
| EMG1D | EMG signal from the non-affected deltoid muscle |
| EMG2D | EMG signal from the affected deltoid muscle |
| OP, PP, SR | Overhead Pumping, Pushdown Pumping, Shoulder Roll |
| $\alpha_x(t), \alpha_y(t), \alpha_z(t)$ | Acceleration components along the $x$-, $y$-, and $z$-axes at time $t$ |
| $\gamma_x(t), \gamma_y(t), \gamma_z(t)$ | Gyroscope components along the $x$-, $y$-, and $z$-axes at time $t$ |
| $\gamma = \sqrt{\gamma_x^2 + \gamma_y^2 + \gamma_z^2}$ | Overall rotational motion magnitude |
| $P(OP), P(PP), P(SR)$ | Aggregated softmax probabilities |
| $T, S$ | Window size and stride step of activity classifier in Section 2.1 |
| $d$ | Input dimension/number of sensor channels in Section 2.1 |

### 2.1 Supervised Learning of Multi-Window Aggregation for Wearable Motion Classification

We develop a supervised learning framework for wearable-device-based activity recognition in which a neural network is trained on motion recordings from a subset of participants and its generalization is evaluated on held-out participants. Suppose the wearable device is equipped with *P* sensing modalities (e.g., accelerometer, gyroscope, magnetometer), where the *p*th modality produces a *d*_*p*_-dimensional measurement at each sample time. The total input dimension (total number of sensor channels) is 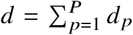. At sample time *t*_i_, we concatenate all modality outputs into a single measurement vector

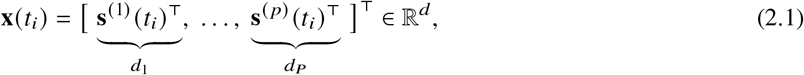

where 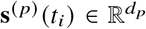 is the output of the *p*th modality. *In this study*, the wearable device is a wrist-worn inertial measurement unit (IMU) with *P* = 2 modalities: a tri-axial accelerometer (**s**^(1)^ ≡ *α, d*_1_ = 3) and a tri-axial gyroscope (**s**^(2)^ ≡ *γ, d*_2_ = 3), giving *d* = 6. This yields a *specific form* of (2.1), as follows

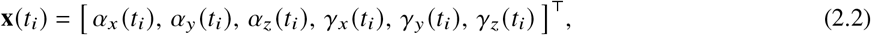

where *α*_*x*_, *α*_*y*_, *α*_*z*_ and *γ*_*x*_, *γ*_*y*_, *γ*_*z*_ denote the accelerometer and gyroscope components along the Cartesian *x*-, *y*-, *z*-axes, respectively. The formulation in (2.1) is kept general so that additional modalities (e.g., magnetometer, barometer, or photoplethysmography) can be incorporated by simply augmenting *P* and expanding the dimension *d* without altering the downstream methodology.

A single motion recording is a time series 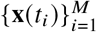, where *M* is the total number of samples in that recording. We denote the sampling frequency of the wearable device by *f*_W_, so that consecutive samples are separated by Δ*t* = 1/ *f*_W_. Suppose there are *C* distinct activity classes *A* = {*a*_1_, …, *a*_*C*_ }. Each recording is collected under a known exercise condition and is therefore associated with a class label *y* ∈ *A*.

Therefore, the general supervised learning task can be formulated as follows: Given a training set 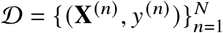, where 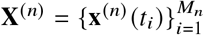 is the *n*th motion recording and *y* ^(*n*)^ ∈ *A* is its associated activity label, the goal of the supervised classification task is to learn a mapping *f*_***θ***_ : χ → *A*, parameterized by *θ*, that assigns the correct activity label to each recording. The parameters are obtained by minimizing the empirical risk

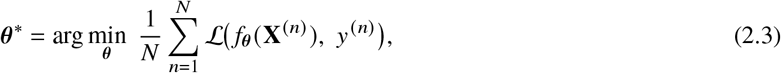

where ℒ is a suitable loss function (e.g., the cross-entropy loss for multi-class classification). At inference time, the predicted label for a new recording **X**^∗^ is given by *ŷ* = *f*_***θ***_ ^∗^ (**X**^∗^).

*In this study*, we focus on the activity set that is consisted of *C* = 3 classes: overhead pumping, pushdown pumping, and shoulder roll, i.e. *A* = {OP, PP, SR}. Because recordings vary in length across trials, we implement *f*_***θ***_ by operating on shorter, fixed-size segments. Each recording is first normalized channel-wise to reduce differences in sensor scale and offset across trials. The normalized signal is then partitioned using a sliding window of length *T* samples with a stride of *S* samples, producing a set of overlapping segments that densely cover the recording. Because every recording is collected under a single known exercise condition, each window inherits the activity label *y* ^(*n*)^ of its parent recording. Let *N*_*w*_ denote the total number of windows extracted across all training recordings. Each window is a *T* × *d* matrix and is passed through a temporal neural network that extracts hierarchical motion features and outputs a softmax score vector over *A*. At inference time, a single recording generally yields multiple overlapping windows. The window-level softmax vectors from the same recording are averaged to produce a recording-level score 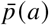 for each class *a* ∈ *A*, and the predicted activity is determined by 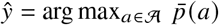. This averaging reduces sensitivity to local temporal variations within a recording and stabilizes the final prediction. The model is trained on recordings from a subset of participants and evaluated on held-out participants who do not appear in the training set, so as to assess generalization to unseen individuals. Results are presented in Section 3.3.

### 2.2 Supervised RNN Inference of Target Signals from Wearable Measurements

Once the activity class *ŷ* has been identified in Stage 1 (Section 2.1), the second stage of the REINA framework uses the same wearable measurements to infer physiological signals that, while clinically informative, are too costly or impractical to acquire continuously outside a laboratory setting.

Let *ξ*(*t*_*i*_) ∈ ℝ^*K*^ denote the target signal vector at sample time *t*_*i*_, where *K* is the number of output channels. The formulation is general: *ξ* may EMG envelopes, joint torques, or any other physiological quantity that was simultaneously recorded during training but will be unavailable at deployment. In this study, the target signal is the preprocessed EMG envelope. Specifically, let *j* ∈ {1, 2 } index the body side (non-affected and affected, respectively) and let *s*_1_, …, *s*_*K*_ index the sensor patches on that side. Then

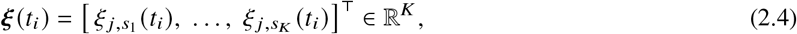

where 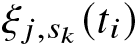 is the preprocessed EMG envelope from the *k*th patch on side *j* . In the present work we focus on the deltoid channel (*K* = 1) on each side (see Table 1), though the formulation applies to arbitrary *K*.

In general the target signal may be sampled at a frequency *f*_T_ that differs from the wearable sampling frequency *f*_W_. To obtain synchronized input–output pairs, we adopt the wearable time axis 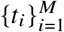 as the reference and resample the target signal onto this grid via linear interpolation, yielding an aligned pair (x(*t*_*i*_),*ξ* (*t*_*i*_)) at every wearable sample instant. In this study *f*_T_ = *f*_EMG_, the sampling frequency of the EMG device.

With the aligned data in hand, the inference task is formulated as a supervised regression problem. Recall from (2.2) that *t*_*i*_ ∈ ℝ^*d*^ is the wearable measurement vector at time x(*t*_*i*_). Because the wearable measurement **x**(*t*_*i*_) captures only a low-dimensional projection of the full musculoskeletal state, the instantaneous recorded data alone does not uniquely determine the corresponding physiological response: biomechanically distinct motor strategies can produce nearly identical kinematic snapshots [9, 10]. In other words, the mapping from observed kinematics to the target signal is non-Markovian with respect to the wearable observations. The short-term movement history, i.e. a short segment of the observed time series, however, provides the temporal memory needed to resolve this ambiguity. Therefore, we equip the model with a short input segment of length *τ*,

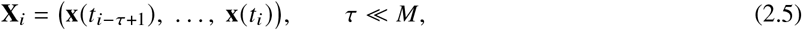

where *τ* is the memory length and *M* is the total number of samples in the recording, as defined in Section 2.1. Then, we seek a parameterized mapping ℱ_*ϕ*_: ℝ ^*τ*×*d*^ → ℝ^*K*^ such that the predicted target

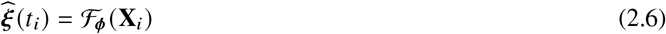

approximates the true target vector *ξ* (*t*_*i*_). Following the same empirical minimization principle as in (2.3), the parameters ***ϕ*** are obtained by minimizing the mean-squared-error loss over all training recordings:

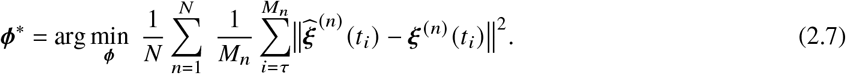

A separate model is trained for each recognized activity class, allowing ℱ_***ϕ***_ to specialize its learned temporal features to the dynamics of a particular exercise. We implement ℱ_***ϕ***_ with a recurrent neural network (RNN). At each time step the hidden state is updated according to

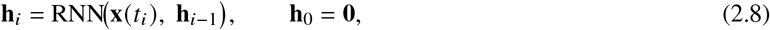

where **h**_*i*_ ∈ ℝ ^*h*^ encodes the temporal information accumulated from the wearable measurements within the fixed time window. The target prediction is then obtained via a learned output layer

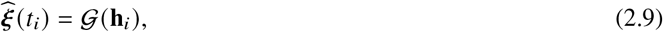

where *G* : ℝ ^*h*^ → ℝ ^*K*^ is a linear output layer. At inference time, given a new wearable recording whose activity has been classified as *ŷ* in Stage 1, the corresponding activity-specific model 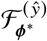 is applied to produce the estimated target trajectory 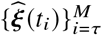. The following Root Mean Square Error (RMSE) can therefore be used to quantify the prediction

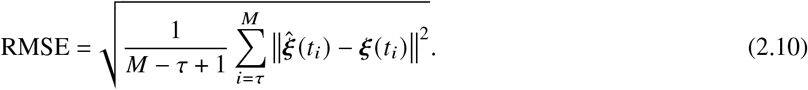

## 3 Data and Results

### 3.1 Data source and preprocessing

#### Data source

Multimodal data were collected from a cohort of breast cancer survivors (*N* = 30). Eligible participants were adults (age ≥ 18 years) who had a history of being diagnosed or treated for breast cancer. Written informed consent was obtained from all participants prior to participation, with clear explanation of study procedures, risks, voluntary participation, and the right to withdraw at any time without penalty. The study protocol was approved by the Institutional Review Board (IRB) of the University of Missouri-Kansas City (IRB Project Number: 2099328).

During the study, each participant performed a set of standardized upper-limb rehabilitation exercises included in The-Optimal-Lymph-Flow (TOLF) intervention program [20, 21, 22, 23], including deep breathing, clasp and spread, horizontal pumping, overhead pumping, pushdown pumping, and shoulder roll. The example videos of the exercises are publicly available on the following website: https://optimallymph.org/instruction-videos.

Each exercise in the set was performed for a duration of 2 minutes, with rest periods in between to minimize fatigue. Each set was repeated 3 to 5 times to ensure data reliability.

During the exercises, each participant was equipped with the Delsys Trigno wireless EMG sensors placed on both upper arms to monitor muscle activity. Additionally, participants wore a Google Pixel 2 smartwatch on their left wrist to capture motion data, including accelerometer and gyroscope readings. Thus, two synchronized data streams were recorded for each participant.

#### Random Selection of Patients

The current study is intended as a methodological demonstration rather than a population-level inferential study. To illustrate the proposed multimodal analytical approach, three representative participants were selected from the larger cohort. Future studies will apply the methodology to the full dataset and evaluate its generalizability across a larger sample. Thus, in the current study, we randomly selected three patients from the study cohort, denoted as Patients I, II, and III, and use their data for investigation.

#### EMG data preprocessing for reference generation

The EMG data used as the *reference* in this study (i.e., the EMG envelope) is processed from the raw signal through band-pass filtering (20–450 Hz [24]), full-wave rectification (i.e., taking the absolute value), and moving root-mean-square (RMS) smoothing. The moving RMS operation computes the signal energy over a sliding window. In this study, a 50 ms window is adopted to reduce excessive high-frequency fluctuations.

##### Remark 1

(Observations on EMG–IMU data correlations). *We showcase the processed EMG signals and corresponding smartwatch measurements for Patient II in Figures 3–5 for the overhead pumping, pushdown pumping, and shoulder roll activities. Qualitative comparison shows that the EMG data and smartwatch signals are consistent, particularly in the temporal alignment of corresponding peaks and troughs. It is also worth noting that, in some cases, the wearable sensor data appear to capture the rehabilitation process more accurately than the EMG measurements. For example, the gyroscope signal in Figure 3 (d) exhibits two repetitions (major impulses) corresponding to the overhead pumping activity performed during the exercise, whereas the corresponding EMG signal does not explicitly exhibit this pattern*.

**Figure 3:**
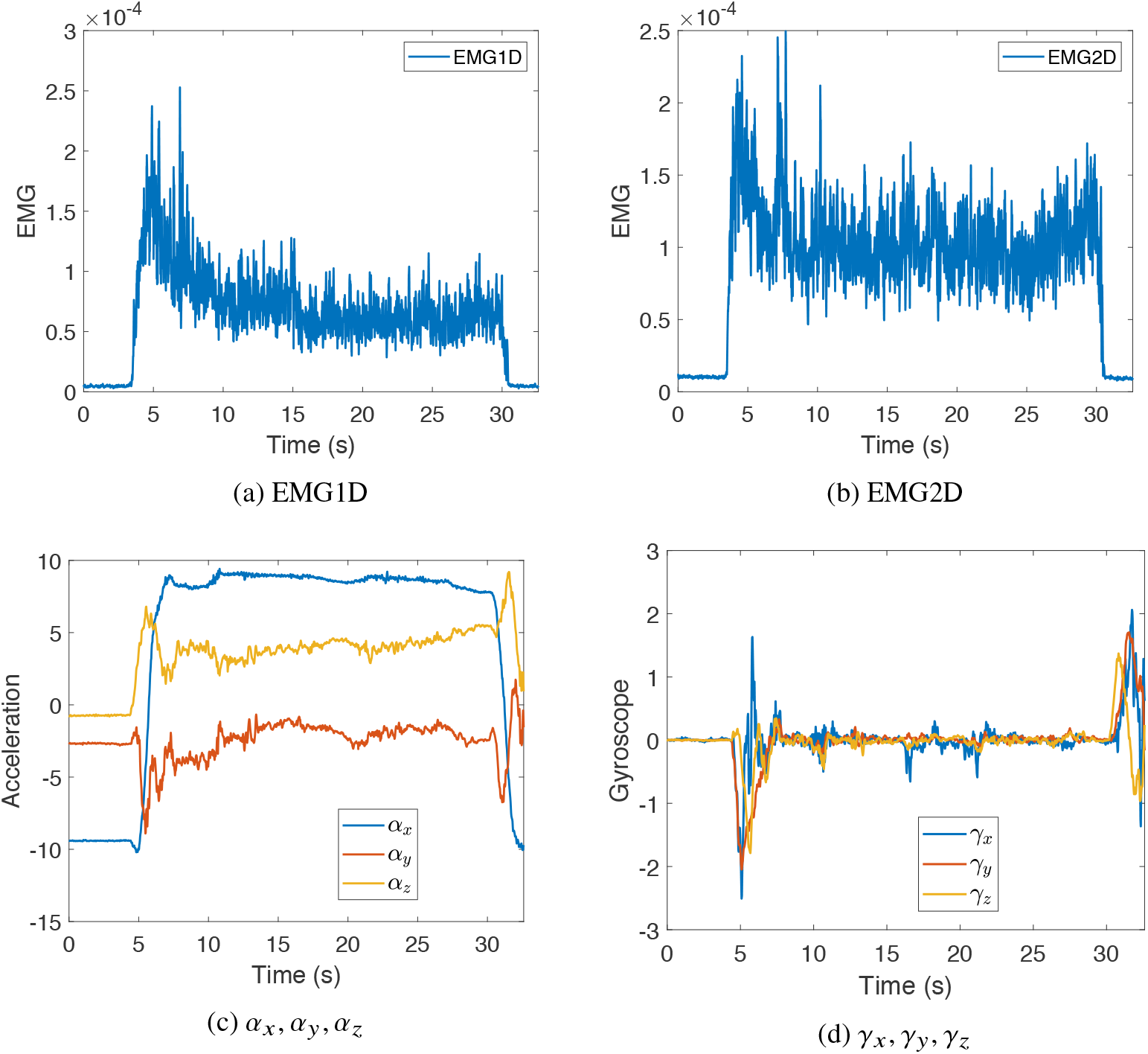
Time series of Patient 2 Overhead Pumping EMG and smartwatch signals.

**Figure 4:**
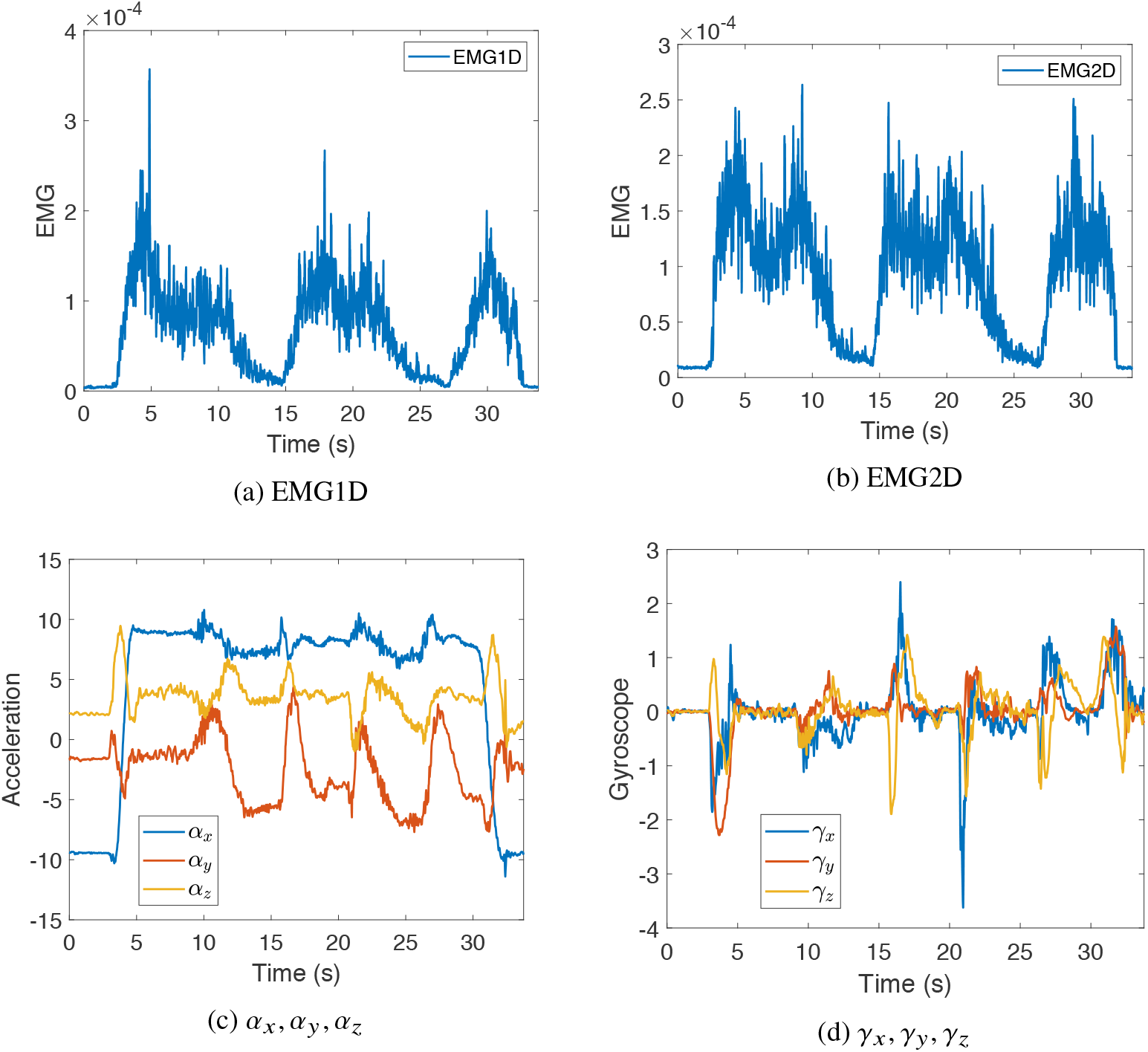
Time series of Patient 2 Pushdown Pumping EMG and smartwatch signals.

**Figure 5:**
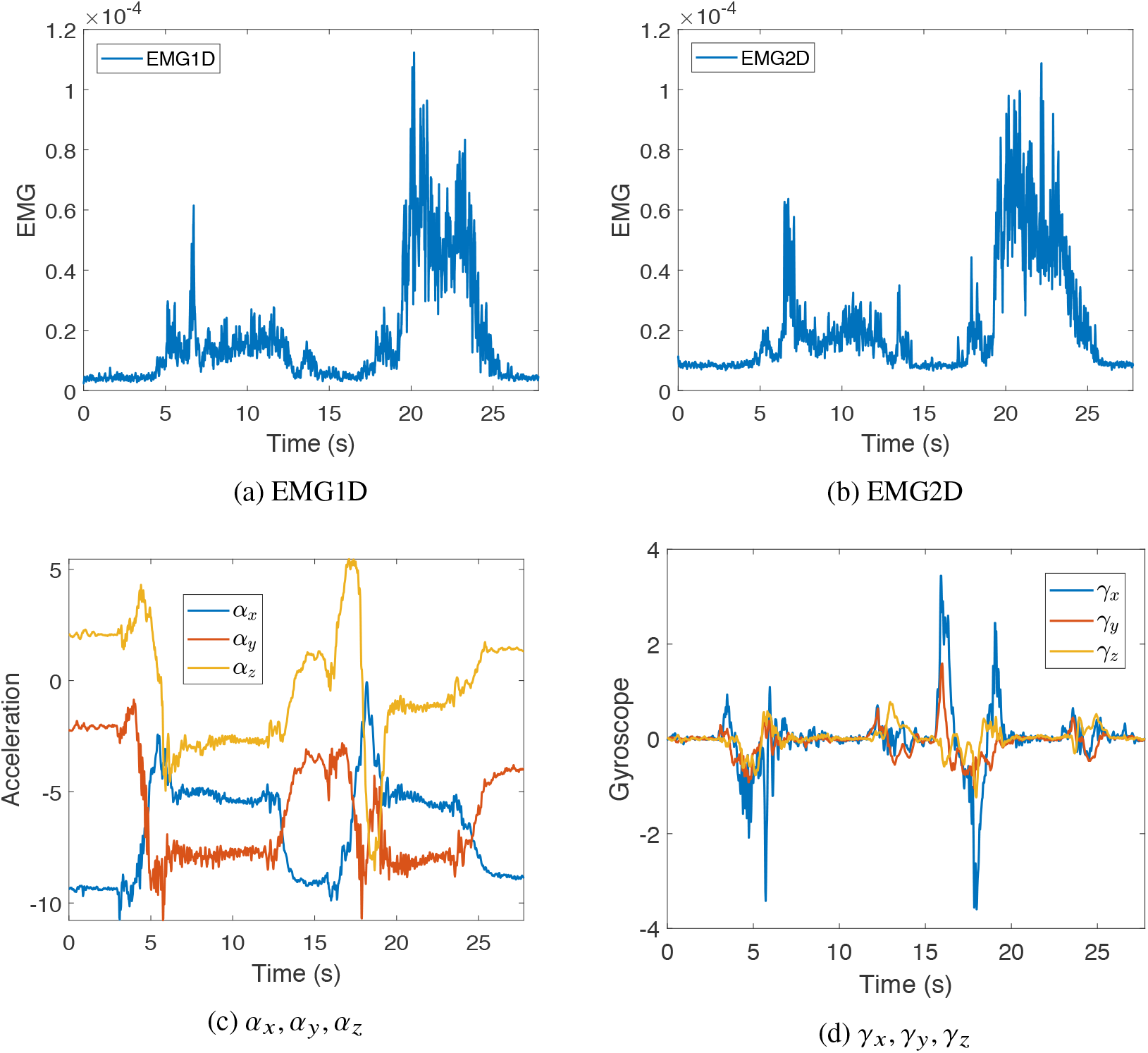
Time series of Patient 2 Shoulder Roll EMG and smartwatch signals.

To quantify the observed EMG-IMU data correlations, we randomly select three datasets for each of the overhead pumping and shoulder roll activities from Patient I. In addition, we randomly select two datasets for each ot these two activities from Patient II. For each patient, the average correlations are reported in Table 2. The table shows the correlations between the EMG signal and the smartwatch measurements, including *α*_*z*_ and 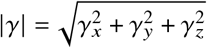, where we recall the definitions of those notations in Table 1. Moderately higher correlations observed in some cases provide empirical support and further strengthen the motivation for inferring EMG signals from smartwatch measurements. We note that some average correlation values are reduced by variability across datasets. For example, for the overhead pumping task of Patient II, one dataset yields an 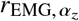 value around 0.6, whereas another yields a value of approximately 0.2. For completeness, the full set of observed correlation results is provided in Table 8.

**Table 2:**
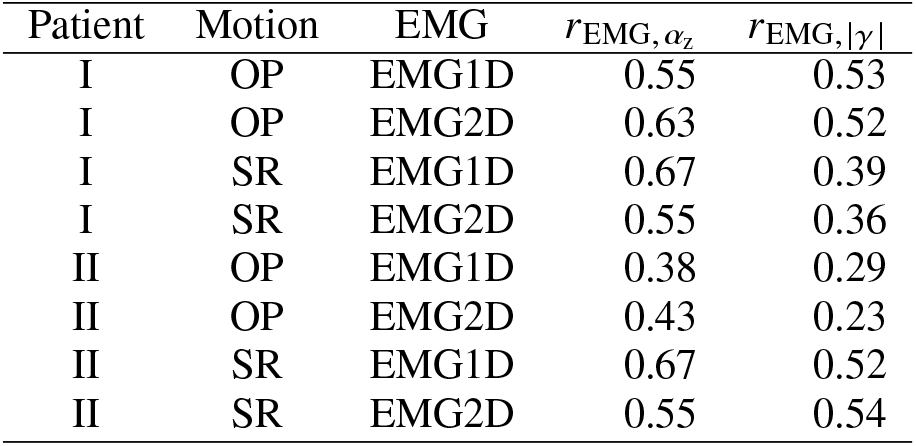
Average EMG–IMU correlations across observations.

### 3.2 Motion trajectory point cloud of Participants

We recorded the motions of TOLF participants from the randomly selected breast cancer cohort (Patients I, II, and III). The acceleration and gyroscope motion trajectories are shown in Figure 6. For each IMU sensor channel signal, denoted by *s* (where *s* = *α*_*X*_, *α*_*y*_, *α*_*z*_, *γ*_*X*_, *γ*_*y*_, *γ*_*z*_), we apply normalization using

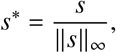

where ||*s* ||_∞_ denotes the maximum absolute value of *s*. Figure 6 illustrates distinct patterns of motion manifolds for different activities (overhead pumping, pushdown pumping, and shoulder roll). These characteristic patterns provide intuition and motivation for designing a machine-learning approach to recognize the participants’ activities from IMU sensor data.

**Figure 6:**
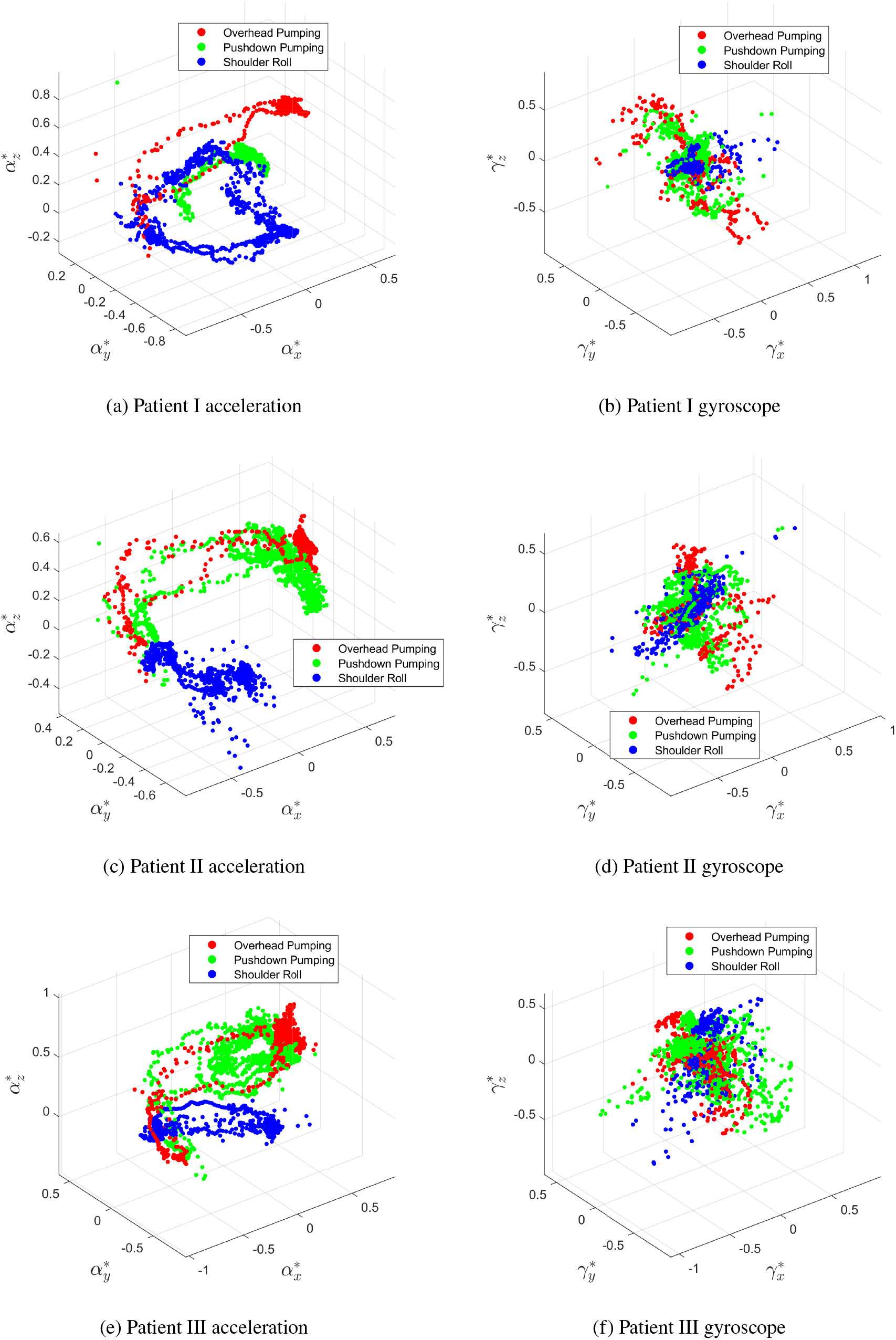
Motion manifold for Overhead Pumping, Pushdown Pumping, and Shoulder Roll.

### 3.3 Activity recognition with supervised learning

First, we specify the parameters used for the methodology described in Section 2.1: *T* = 200 samples (i.e., each window spans 4 seconds of the signal at a sampling rate of 50 Hz), with a sample stride of S=25. The dimension of sensor channels is *d* = 6 as specified in (2.2). *N*_*w*_ is recording-dependent and related to *T* and *S* by its definition.

We then evaluate the efficacy of the supervised learning model with multiple-window aggregation introduced in Section 2.1 using a rotational training–testing scheme across three participants. In each rotation, the model is trained on data from two participants across multiple recordings and evaluated on the remaining participant. To be specific, we test the following three cases:

**Case (i)**: train on data from Patients I and III, and evaluate on Patient II;

**Case (ii)**: train on data from Patients I and II, and evaluate on Patient III;

**Case (iii)**: train on data from Patients II and III, and evaluate on Patient I.

We recall that *P*(OP), *P*(PP), and *P*(SR) (defined in Table 1) denote aggregated softmax probabilities, computed by averaging the window-wise softmax outputs across all *N*_*w*_ windows of a recording. We define the activity prediction *confidence* as follows:

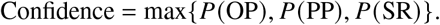

The results are summarized in Tables 3, 4, and 5, for Cases (i), (ii), and (iii), respectively. “ID” in the tables means the label of recordings. The tables show that the supervised multi-window neural network model generally provides accurate classification predictions. In most cases, the activities of the participants are correctly identified. One exception occurs in Case (iii), highlighted in Table 5, where PP (Recording A) is misclassified as OP. However, the predicted probabilities for OP and PP are 0.587 and 0.410, respectively, indicating a relatively small margin. This suggests that the model does not make a decisive misprediction. Rather, for this particular sample, the comparable probabilities suggest that the model’s assessment of this sample remains reasonable, even though the prediction result is incorrect. Nevertheless, the supervised network model achieves overall accurate and robust activity recognition on the test samples.

**Table 3:** Results of activity classification prediction for Case (i).

| True | ID | N | Prediction | Confidence | P(OP) | P(PP) | P(SR) | Correct |
| --- | --- | --- | --- | --- | --- | --- | --- | --- |
| OP | A | 66 | OP | 0.670 | 0.670 | 0.277 | 0.053 | Y |
| OP | B | 70 | OP | 0.656 | 0.656 | 0.257 | 0.087 | Y |
| OP | C | 59 | OP | 0.654 | 0.654 | 0.321 | 0.025 | Y |
| PP | A | 65 | PP | 0.831 | 0.143 | 0.831 | 0.027 | Y |
| PP | B | 70 | PP | 0.869 | 0.128 | 0.869 | 0.003 | Y |
| PP | C | 70 | PP | 0.750 | 0.132 | 0.750 | 0.118 | Y |
| SR | A | 62 | SR | 0.884 | 0.012 | 0.104 | 0.884 | Y |
| SR | B | 61 | SR | 0.986 | 0.004 | 0.011 | 0.986 | Y |

**Table 4:** Results of activity classification prediction for Case (ii).

| True | ID | $N_w$ | Prediction | Confidence | P(OP) | P(PP) | P(SR) | Correct |
| --- | --- | --- | --- | --- | --- | --- | --- | --- |
| OP | A | 63 | OP | 0.739 | 0.739 | 0.258 | 0.003 | Y |
| OP | B | 59 | OP | 0.810 | 0.810 | 0.186 | 0.005 | Y |
| OP | C | 58 | OP | 0.688 | 0.688 | 0.206 | 0.106 | Y |
| PP | A | 60 | PP | 0.796 | 0.194 | 0.796 | 0.009 | Y |
| PP | B | 63 | PP | 0.875 | 0.103 | 0.875 | 0.022 | Y |
| PP | C | 46 | PP | 0.905 | 0.004 | 0.905 | 0.091 | Y |
| SR | A | 42 | SR | 0.996 | 0.002 | 0.003 | 0.996 | Y |
| SR | B | 50 | SR | 0.786 | 0.133 | 0.081 | 0.786 | Y |
| SR | C | 44 | SR | 0.522 | 0.154 | 0.324 | 0.522 | Y |

**Table 5:** Results of activity classification prediction for Case (iii).

| True | ID | $N_w$ | Prediction | Confidence | P(OP) | P(PP) | P(SR) | Correct |
| --- | --- | --- | --- | --- | --- | --- | --- | --- |
| OP | A | 71 | OP | 0.999 | 0.999 | 0.001 | 0.000 | Y |
| OP | B | 59 | OP | 0.994 | 0.994 | 0.006 | 0.000 | Y |
| OP | C | 71 | OP | 0.999 | 0.999 | 0.001 | 0.000 | Y |
| OP | D | 54 | OP | 0.924 | 0.924 | 0.073 | 0.003 | Y |
| **PP** | A | 75 | **OP** | 0.587 | 0.587 | 0.410 | 0.002 | N |
| PP | B | 64 | PP | 0.645 | 0.347 | 0.645 | 0.008 | Y |
| SR | A | 47 | SR | 0.778 | 0.041 | 0.181 | 0.778 | Y |
| SR | B | 55 | SR | 0.960 | 0.002 | 0.039 | 0.960 | Y |
| SR | C | 47 | SR | 0.778 | 0.041 | 0.181 | 0.778 | Y |

### 3.4 RNN predictions for EMG signals

We employ the RNN framework described in Section 2.2 to predict EMG signals, focusing on EMG1D and EMG2D (see Table 1 for definitions). Specifically, we consider overhead pumping and shoulder roll for Patient I; pushdown pumping and shoulder roll for Patient II; and overhead pumping and pushdown pumping for Patient III. The reference EMG data are generated from the raw recordings using the preprocessing procedure in Section 3.1. Comparisons between predicted and reference EMG envelopes are presented in Figures 7, 8, and 9. The results demonstrate close agreement between predictions and the reference EMG across motions and participants. The recording IDs used to generate the reference EMG data are specified in the corresponding figure sub-captions. To further quantify performance, we present the RMSE (defined in (2.10)) of the RNN predictions in Table 6, where RMSE1 is for EMG1D; RMSE2 is for EMG2D. Those results indicate accurate prediction. We also compute the correlation coefficients between predicted and reference EMG, summarized in Table 7. Here, *r*_EMG1D_ and *r*_EMG2D_ denote the correlations for EMG1D and EMG2D, respectively. The consistently high correlations confirm that the RNN framework effectively captures the temporal dynamics of the EMG signals across different motions and participants.

**Table 6:** RMSE (defined in (2.10)) of EMG predictions relative to the reference signals (EMG1D and EMG2D) for the tested patients across different activities.

| Patient and activity | RMSE1 | RMSE2 |
| --- | --- | --- |
| Patient I: OP | 2.546e-05 | 5.889e-05 |
| Patient I: SR | 7.963e-06 | 8.422e-06 |
| Patient II: SR | 7.401e-06 | 7.990e-06 |
| Patient II: PP | 2.336e-05 | 2.663e-05 |
| Patient III: OP | 5.094e-05 | 4.571e-05 |
| Patient III: PP | 3.733e-05 | 3.969e-05 |

**Table 7:** Correlation between reference and RNN-predicted reference EMG.

| Participant | Motion | ID | $r_{\text{EMG1D}}$ | $r_{\text{EMG2D}}$ |
| --- | --- | --- | --- | --- |
| Patient I | OP | B | 0.8114 | 0.8347 |
| Patient I | SR | B | 0.7789 | 0.7196 |
| Patient II | PP | B | 0.8879 | 0.8856 |
| Patient II | SR | B | 0.9234 | 0.9145 |
| Patient III | OP | C | 0.7831 | 0.7806 |
| Patient III | PP | B | 0.8817 | 0.8349 |

**Table 8:** EMG–IMU correlations across observations.

| Patient | Motion | EMG | Dataset | $r_{\text{EMG}, \alpha_\gamma}$ | $r_{\text{EMG}, \vert \gamma\vert }$ |
| --- | --- | --- | --- | --- | --- |
| I | OP | EMG1D | A | 0.53 | 0.48 |
| I | OP | EMG2D | A | 0.64 | 0.47 |
| I | OP | EMG1D | B | 0.60 | 0.63 |
| I | OP | EMG2D | B | 0.60 | 0.61 |
| I | OP | EMG1D | C | 0.53 | 0.48 |
| I | OP | EMG2D | C | 0.64 | 0.47 |
| I | SR | EMG1D | A | 0.66 | 0.37 |
| I | SR | EMG2D | A | 0.55 | 0.35 |
| I | SR | EMG1D | B | 0.70 | 0.42 |
| I | SR | EMG2D | B | 0.56 | 0.37 |
| I | SR | EMG1D | C | 0.66 | 0.37 |
| I | SR | EMG2D | C | 0.55 | 0.35 |
| II | SR | EMG1D | A | 0.41 | 0.59 |
| II | SR | EMG2D | A | 0.61 | 0.56 |
| II | SR | EMG1D | B | 0.64 | 0.44 |
| II | SR | EMG2D | B | 0.61 | 0.52 |
| II | OP | EMG1D | A | 0.18 | 0.29 |
| II | OP | EMG2D | A | 0.20 | 0.23 |
| II | OP | EMG1D | B | 0.55 | 0.29 |
| II | OP | EMG2D | B | 0.65 | 0.23 |

**Figure 7:**
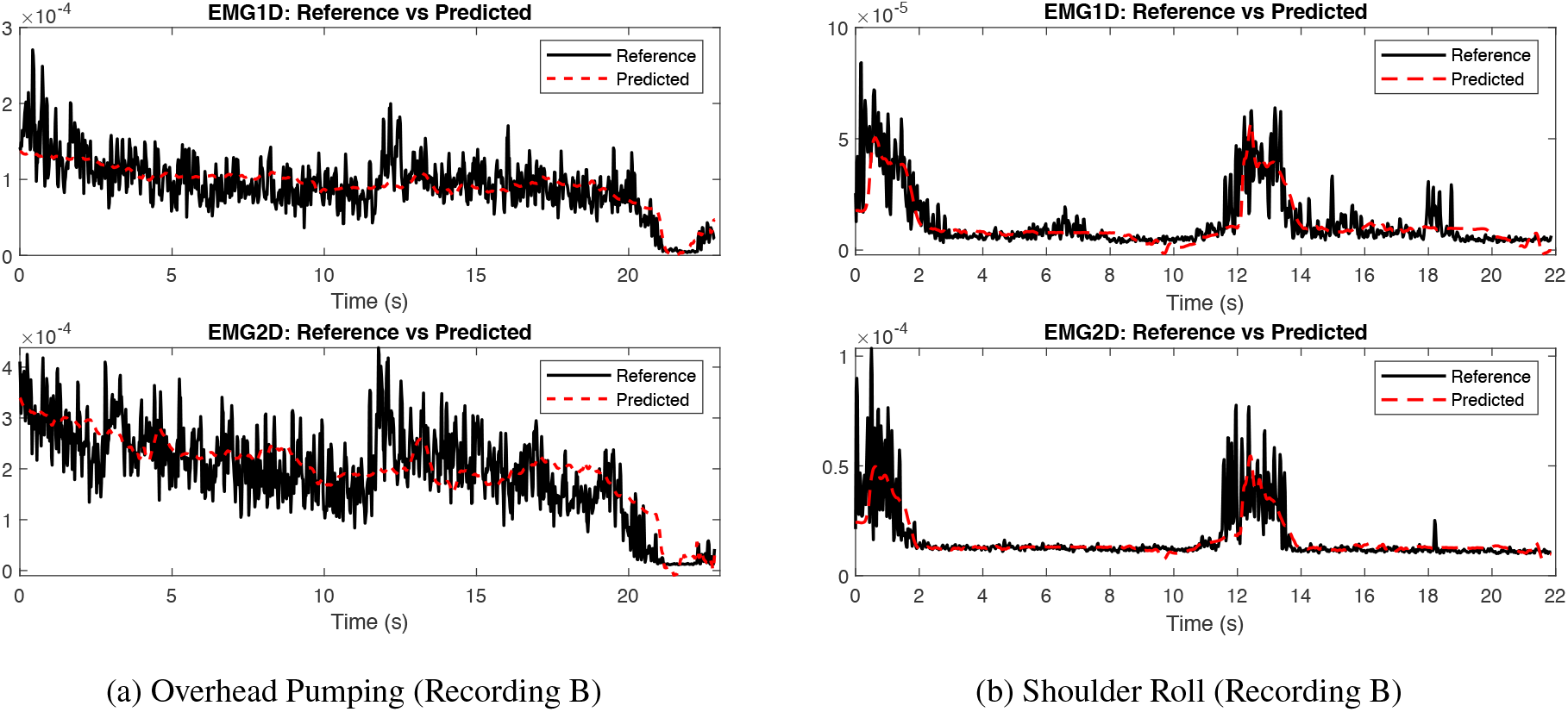
Comparison of the reference EMG and the RNN prediction for Patient I.

**Figure 8:**
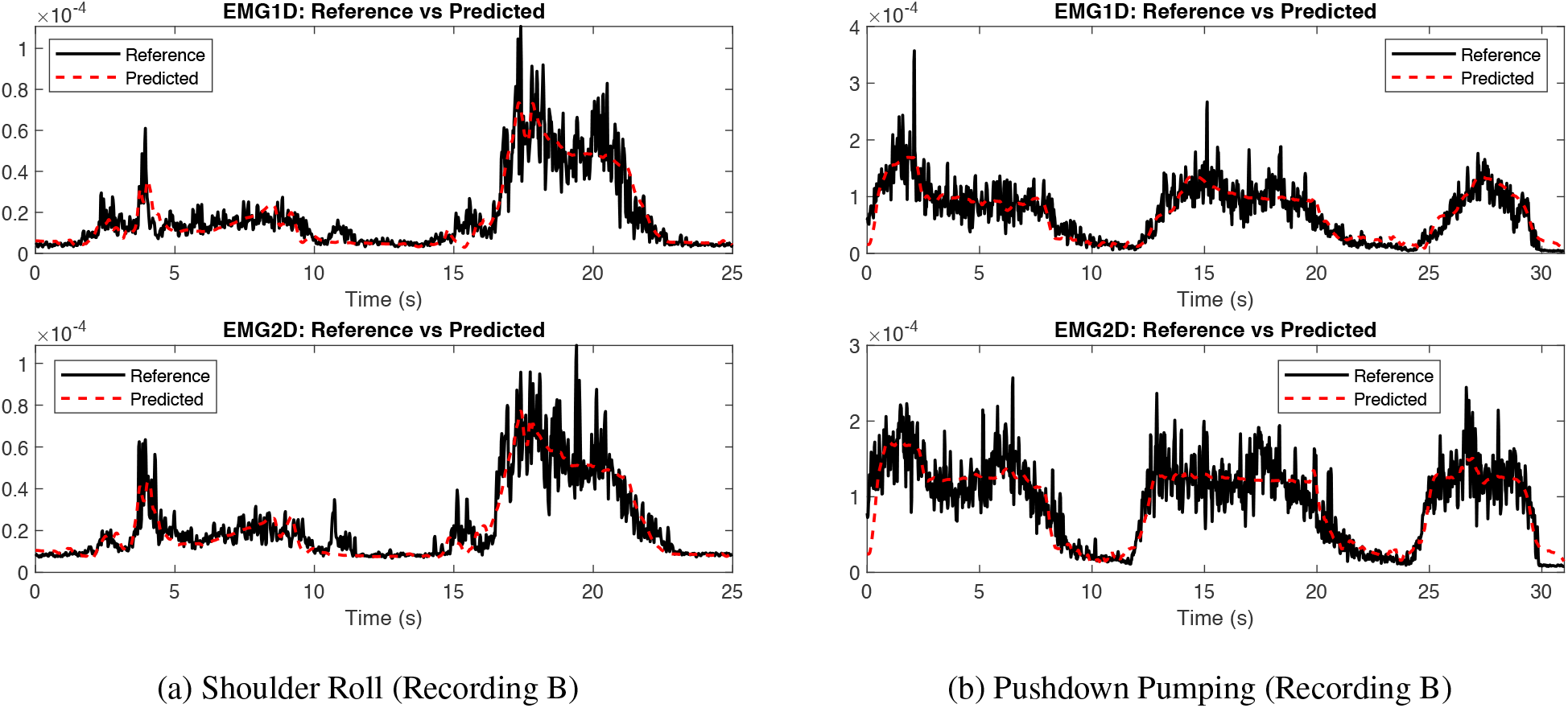
Comparison of the reference EMG and the RNN prediction for Patient II.

**Figure 9:**
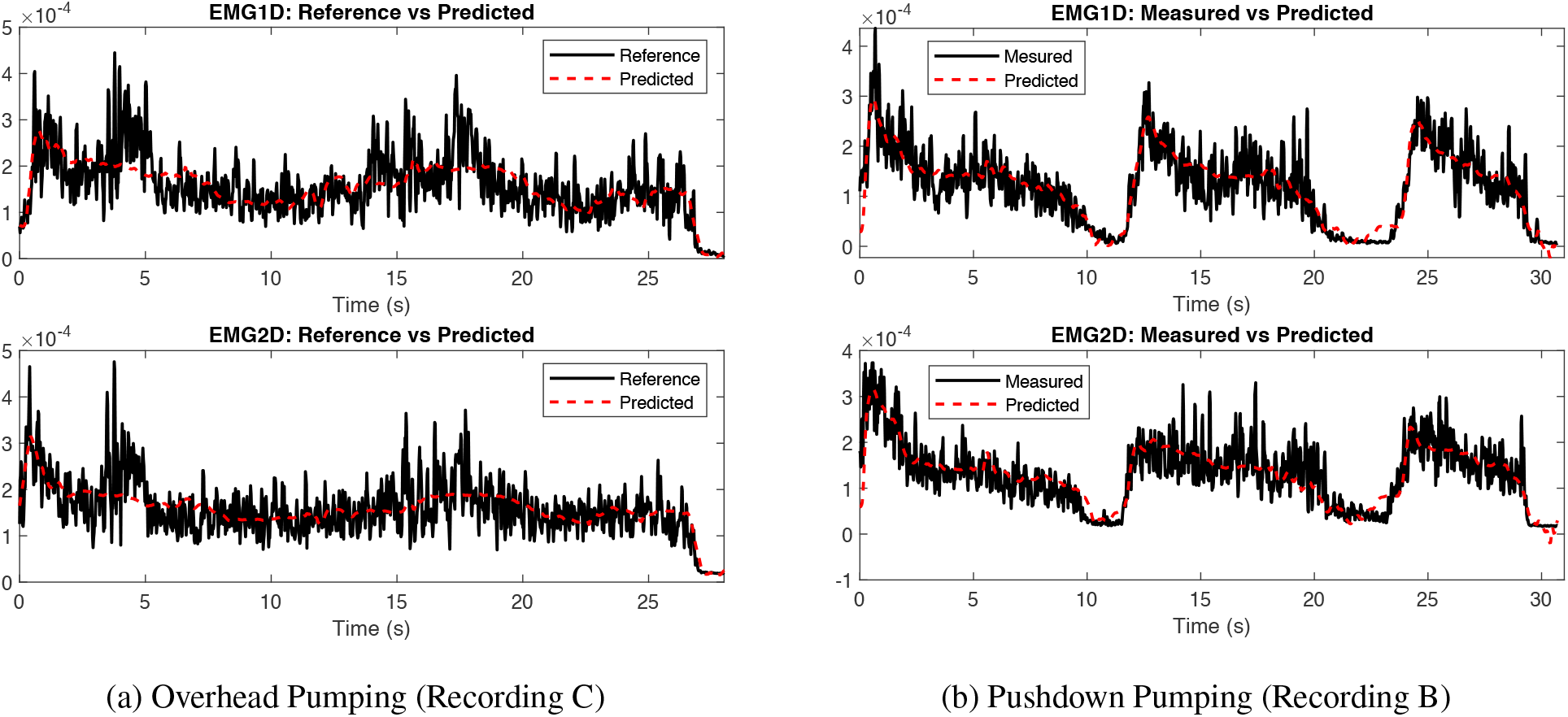
Comparison of the reference EMG and the RNN prediction for Patient III.

## 4 Conclusion and Future Work

This work has proposed REINA, a two-stage deep-learning framework that integrates activity recognition (with supervised learning) and EMG inference (with RNN) from smartwatch IMU signals for remote monitoring of breast cancer rehabilitation. The results demonstrate that REINA can successfully recognize rehabilitation activities and then accurately infer EMG signals from wearable IMU measurements. The proposed framework provides a reliable, real-time, and cost-effective approach for monitoring breast cancer rehabilitation.

The present study has several limitations. The current work primarily serves as a proof-of-concept study based on a relatively small cohort and a modest set of rehabilitation activities. In addition, the analysis focuses mainly on deltoid EMG signals during representative rehabilitation motions. Nevertheless, the results demonstrate the feasibility of activity recognition and EMG inference from wearable IMU measurements using REINA.

Future work will investigate larger and more diverse rehabilitation datasets involving broader participant populations and rehabilitation activities. Additional muscle groups and more complex daily-life motions will also be explored to further evaluate the generalizability and clinical applicability of the proposed framework. Individualized and more adaptive learning strategies, multimodal wearable sensing, and more advanced temporal learning architectures may further improve the robustness and efficiency of wearable-based EMG inference.

## Data Availability

The participant-level data are not publicly available because they contain sensitive human-subject information and are subject to participant-privacy protections and the conditions of the IRB-approved protocol.

## Acknowledgements

This study was supported by The UMKC (University of Missouri-Kansas City) and the Kauffman Foundation – Entrepreneurship Innovation Grant (KHG30) and by The UMKC Research Advisory Council Tier 1 Research Award (K9022) with Dr. Mei R Fu as the PI and Dr. Bowen Liu and Dr. Gregory King as the Co-PIs. The content is solely the responsibility of the authors and does not necessarily represent the official views of the funder.

**Appendix A Observations on EMG-IMU Data Correlations**

